# Emerging costs in a “hidden” workforce: The longitudinal psychosocial effects of caregiving during the COVID-19 pandemic among Norwegian adults

**DOI:** 10.1101/2023.06.06.23290515

**Authors:** Thomas Hansen, Kamila Hynek, Anne McMunn, Ragnhild Bang Nes, Vegard Skirbekk, Margarethe Vollrath, Fredrik Methi

## Abstract

During COVID-19 many informal caregivers experienced increased caregiving load while access to formal and informal support systems and coping resources decreased. Little is known about the psychosocial costs of these challenges for an essential yet vulnerable and “hidden” frontline workforce. This study explores and compares changes in psychosocial well-being (psychological well-being, psychological ill-being, and loneliness) before and across up to three stages of the COVID-19 pandemic among caregivers and non-caregivers. We also examine predictors of psychosocial well-being among caregivers during the peak of the pandemic. We use longitudinal data collected online in the Norwegian Counties Public Health Survey (age 18–92) in four countries and up to four data points (n=14,881). Caregivers are those who provide care unpaid, continuous (≥ monthly across all time points) help to someone with health problems. Findings show that levels of psychosocial well-being first remained stable but later, during the peak stages of the pandemic, dropped markedly. Caregivers (13−15% of the samples) report lower psychosocial well-being than non-caregivers both before and during the pandemic. Caregivers seem especially vulnerable in terms of ill-being, and during the peak of the pandemic caregivers report higher net levels of worry (OR = 1.22, p< .01) and anxiety (OR = 1.23, p< .01) than non-caregivers. As expected, impacts are graver for caregivers who provide more intensive care and those reporting health problems or poor access to social support. Our study findings are valuable information for interventions to support caregivers during this and future pandemics.

## 1. Introduction

The COVID-19 pandemic has generated immense challenges for health care systems worldwide (Abbas 2021; Eurofound 2022). One particularly relevant group for these systems has not received much research attention, namely informal caregivers, who provide unpaid care to individuals with long-term illnesses or other health-related needs. Considered key partners in disease management and care coordination, informal caregivers account for about half of the total care provided in Norway (Opinion 2021). Even in normal times, evidence suggests that caregivers experience burden and distress that threatens their health and well-being, and in turn, their ability to care for their care recipients (Adelman et al. 2014; Hansen and Slagsvold, 2012). However, there is widespread concern among caregiver advocacy groups and others that the pandemic has created new and unique challenges for this vulnerable and “hidden” frontline workforce (Eurocarers 2021).

There are multiple reasons why caregivers may have experienced higher than usual burden and stress during COVID-19. First, COVID-19 has posed particular threats for vulnerable groups, which may have led to increased worry for the care recipient’s health and increased self-isolation and even reluctance to receive care services or admittance to care facilities due to fears about infection or prolonged separation (Onwumere et al. 2021). This isolation may also include exposure to challenging behavioral problems, for example in adults with dementia or drug-related disorders. Second, many caregivers experienced that much needed services (e.g., respite care and day centers) were severely restricted or closed during periods of high infection rates and lockdown restrictions. Third, many caregivers were also left without access to their usual support systems (e.g., friends and colleagues, volunteer support), community-based resources (e.g., cafes and swimming pools), and leisure activities (e.g., choir or exercise groups) (Lightfoot et al. 2021; Onwumere et al. 2021). In sum, the combination of increased caregiver load and reduced access to regular coping resources and respite opportunities may have subjected carers to intense levels of stress during COVID-19.

An emerging literature has begun to document pandemic-related changes in caregiver burden and distress. These studies are mostly based on data from the beginning of the pandemic, small convenience samples, and cross-sectional retrospective self-reports. First, there is Norwegian and international evidence that caregivers, especially women, reported increased caregiving time and intensity during the pandemic (Eurofound 2022; Opinion 2021; Truskinovsky et al. 2022; Zwar et al. 2021). For example, a pan-European study of long-term carers shows that caregiving time increased in all countries, and that women increased their mean weekly hours of care (from 48 to 57) more than men (from 39 to 45) (Eurocarers 2021). This study also shows that most caregivers report that the pandemic negatively affected their social participation (79%), well-being (77%), mental health (67%), access to health services for their care recipient (60%), and their care recipient’s health (54%), with the impacts more severe for female than for male caregivers (ibid.). Similarly, several studies explore retrospective changes in stress, exhaustion, and mental health among caregivers and find increasing problems from before to during the pandemic especially among women (Altieri and Santangelo 2021; Canevelli et al, 2020; Cohen et al. 2021; Park 2021; Truskinovsky et al. 2022). Other studies find that care disruptions and caregiving load increased overall during the pandemic, with these changes associated with worse mental health and well-being (Leggett et al. 2022; Truskinovsky et al. 2022). These reports may be subject to recall bias, and without a comparison group of non-caregivers it is uncertain whether caregivers were affected differently from non-caregivers by the pandemic. In a rare study using panel data, from before and during the beginning of the pandemic, caregivers reported higher psychological distress than non-caregivers at both time points, yet both groups reported about equal absolute levels of increase in distress (Gallagher and Wetherell 2020).

This backdrop highlights the need for population-based prospective studies of caregiver distress, over the longer term and during different phases of the pandemic. Importantly, we lack knowledge about how the population reacted to the second (during the fall of 2020) and later waves of the pandemic, when Norway and many other countries witnessed a dramatic increase in infection rates and issued stronger infection control measures (Nørgaard et al. 2021). While caregivers are a heterogenous group with varied risk profiles and thus likely to react differently to COVID-19, there has been little attention to subgroup differences and risk factors. For example, the research on gender differences is still sparse and needs further investigation. While early evidence suggested that women caregivers were disproportionately affected, this expectation is not a given, as women may have more access to social support and higher resilience and coping ability as caregivers than men (Cohen et al. 2021; Gaugler et al. 2007; Merlani et al. 2011). Furthermore, as most studies either focus on older adults or fail to stratify by age, little is known about age-differences in the reactions and especially the impacts among young carers (for an exception, see Blake-Holmes and McGowan 2022). Moreover, the reactions may vary according to access to social, socioeconomic, and other relevant resources. More adverse impacts are likely—for example, among those who are less socially connected or have compromised health themselves, as has been shown during “normal” times (Hansen and Slagsvold 2015). Finally, extant studies are confined to a few countries and there is little evidence from the Nordic countries. Pandemic-related impacts of caregiving may differ across countries due to an interplay between COVID-19 restrictions and cultural and institutional frameworks. Norway is characterized by relatively comprehensive formal care services (Colombo 2011; Hansen et al. 2013), high levels of gender equality, fairly good health among older adults (Skirbekk et al. 2022), and low pandemic-related infection and mortality rates. These patterns likely mitigate risks for female caregivers in particular. Conversely, the pandemic-related restrictions could lead to a drastic change of habitual arrangements and thus cause immense distress to caregivers usually relying on formal structures to support them, especially regarding the more intimate and comprehensive personal care tasks (Daatland et al. 2011).

This study extends prior work by examining gender-stratified longitudinal change in psychological and social (i.e., psychosocial) well-being by caregiver status in a large probability-based sample of adults. These participants were surveyed before and up to three times during the pandemic, including periods with high infection rates (autumns of 2020 and 2021), periods that show as the peaks of the pandemic (The Norwegian Government 2022; WHO 2022). To be able to see the full impact of the pandemic on the informal care situation, research conducted during its peak is needed (Zwar et al. 2021). To identify heterogeneity in the impacts of the COVID-19 pandemic among caregivers and under-supported groups in need of more attention in future pandemics, we investigate predictors of caregivers’ psychosocial well-being at the peaks of the pandemic. These predictors include sociodemographic factors, health variables, and care-related factors such as frequency of caregiving and the experience of added caregiving load during the pandemic.

To our knowledge, this is the first study of its kind to describe levels and changes in psychosocial well-being of informal caregivers during COVID-19. Clarifying how the intensity and psychosocial costs of informal caregiving have changed during COVID-19 is a critical step toward building the case for increasing public health surveillance and enhancing formal support for this vulnerable yet invaluable workforce (Kent et al. 2020).

## 2. Methods

### 2.1 Data

We use data from the Norwegian Counties Public Health Survey (NCPHS), an online cross-sectional study of a probability sample of community-dwelling individuals aged 18+ (Hansen et al. 2021). In response to the COVID-19 outbreak, two counties (Agder and Nordland), in which data were collected just prior to the outbreak, were selected for a COVID follow-up survey. Altogether, data collections were fielded four times. Pre-pandemic data (t1) was collected in Agder 23 Sept−18 Oct 2019 (*N* = 28,015, response rate (*RR*) = 46%) and in Nordland 27 Jan−16 Feb 2020 (*N* = 24,199, *RR* = 47%). A random sample of 20,196 individuals from these counties was invited to participate in three follow-ups, during 4−18 June 2020 (t2; *N* = 11,953, *RR* = 59%), 18 Nov−4 Dec 2020 (t3; *N* = 11,029, *RR* = 55%), and 6−20 Dec 2021 (t4; *N* = 10,220, *RR* = 52%).

In addition, we use data from two counties (Oslo and Vestland) that were invited to participate in the COVID survey at t3 (*N* = 15,134, *RR* = 39%) and t4 (*N* = 12,588, *RR* = 33%), for which we lack pre-pandemic data. These two counties are more urban (and include Norway’s two largest cities: Oslo and Bergen). They were also more heavily hit by the pandemic and issued stronger public restrictions than Agder and Nordland (the location can be seen in Figure 1). After listwise deletion, the four-counties panel sample comprises data from 14,881 individuals.

**Fig 1.**
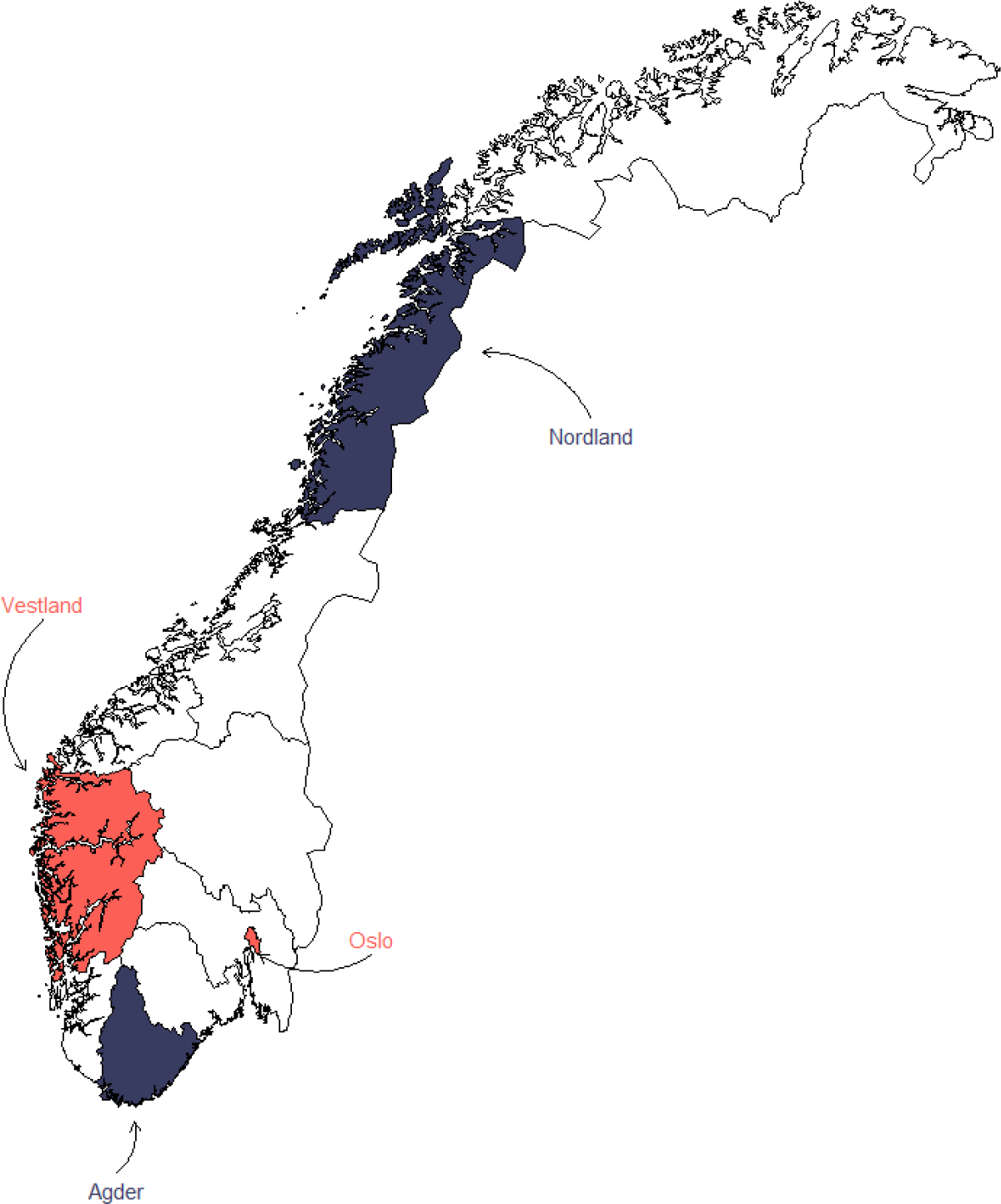
Map of included counties. Counties with two (red) or four (blue) assessments

### 2.2 Caregiving variables

Information about caregiving was included only in the last (t4) data collection, and we thus asked retrospectively about caregiving around the time of the previous data collections. *Caregiver status* questions were introduced by the following question: “Did you, during the whole or part of the period since March 2020, provide regular unpaid help or supervision to someone in need of help due to health problems or old age (e.g., housework, personal care, or supervision)? Please disregard work through a volunteer organization.” Response categories were “no”, “yes, to person(s) in the household”, and “yes, to person(s) outside of the household”. Caregivers (both resident and non-resident) were probed about the *frequency of caregiving* (“How often do/did you provide such help”) with reference to currently and at the time of t1 through t3^1^, and with five response categories (“daily”, “weekly or more often”, “monthly or more often”, “less than monthly”, and “not at this time”). We categorize caregivers as those who provide care at least “monthly or more often” across all available time points.

*Extra caring* (and its appraisal in terms of added stress) was assessed by: “Did your caregiving load increase due to changes in the health- and social services (e.g., home services, respite care, or day care centers) during the fall 2020 lockdown.” Response categories include “No”, “Yes, and it was challenging for me”, and “Yes, but I could handle it”.

### 2.3 Well-being variables

The NCHPS includes a range of indicators of psychological and social well-being. Most are measured by a list of items measuring emotions: “Think about the past 7 days, to what degree did you feel ?” on a scale from 0 (not at all) to 10 (very much). The response format and selection of items conform to conventions and OECD-guidelines in the subjective well-being literature (Nes et al. 2018; OECD 2013).

Psychological well-being refers to how people experience and evaluate their lives, i.e., their emotional and evaluative well-being (Diener 2012). Based on the above items we have constructed an index termed *psychological ill-being*, measured by the mean of three items: *worried*, *anxious*, and *down or sad* (α = .74). Based on the mean of the item *happy* and a single 0−10 *life satisfaction* question (r = .80), we constructed the index *psychological well-being*.

Social well-being can be defined as an appraisal of one’s social interaction and social relationships, and loneliness is one of its core indicators (Aartsen and Hansen 2020). *Loneliness* is measured with a single item that asks about the degree to which one has felt “lonely” (0−10).

### 2.4 Controls

Demographic variables include *gender*, *age* (measured in 10-year intervals), *education* (tertiary (college/university or compulsory/high-school/tertiary) = 1, otherwise non-tertiary 0), *partner status* (married/cohabiting or in a relationship = 1, otherwise 0), *employment status* (full/part time, self-employed, or sickness leave = 1, otherwise 0). *Self-rated health* is measured by a single item recoded into poor (1−2), fair (3), and good (4−5). *Social support* is measured with the 3-item (e.g., “How many people are you so close to that you can count on them if you have great personal problems”) Oslo Support Scale (OSS-3) (α = .60) (Meltzer, 2003). Scores are categorized into poor (score 3−8), moderate (9−11), and strong (12−14) (Bøen et al. 2012). All independent variables are measured at t1 for Agder/Nordland, and t3 for Oslo/Vestland.

### 2.5 Analytical strategy

We conducted two sets of analyses. First, we describe and compare change in well-being variables for caregivers and non-caregiver at the time of the data collections, adjusting for controls. Hence, for Agder/Nordland we use data from before and at three stages of the pandemic (t1 through t4). For Oslo/Vestland we lack pre-pandemic data and use data from late 2020 and late 2021 (the timing of t3 and t4 for Agder/Nordland). To shed additional light on the substantive importance of the observed changes (i.e., how many are “suffering”?), we also show rates of “low” well-being across the three time points. “Low” refers to scores at the undesirable end of the scales, i.e. scores ≥6 for negatively worded items (e.g., lonely) and ≤4 for positively worded items (e.g., happy). Second, we concentrate on caregivers and examine predictors of well-being variables. We analyze dependent variables both cross-sectionally (at t3/t4) and longitudinally (at t3/t4 with control for respective dependent variables at t1). We focus on three sets of predictors: (i) background factors (age, marital status, education, employment status, and health), (ii) caregiving factors (resident vs. non-resident caregiving, frequency of caregiving), and (iii) added caregiving load during the pandemic. All analyses are stratified by gender and performed using Stata v.15.

## 3. Results

### 3.1 Descriptive statistics

About 15% in Agder/Nordland and 13% in Oslo/Vestland provide care monthly or more often during all available time points. Respectively 77% and 80% in these regions do not provide any care at any time point (excluded from the analysis are the 8% and 7%, respectively, that provided care on at least one time point but not monthly or more often throughout). There is significant within-person consistency and change in the frequency of caregiving over time (see transition plot in supplementary online resource Figure S1). Across data collections, among those defined as caregivers (≥monthly in all waves), 29−36% reported to provide care monthly, 53−57% weekly, and 11−16% daily. We observe a trend towards increasing frequency among the caregivers, and that more individuals enter than exit the caregiver role during the pandemic.

We also find that most (70.6%) caregivers cared for someone outside of the household, while 29.4% cared for a person in their household. Furthermore, a minority of caregivers (27.2%) reported that their caregiving load increased during lockdown. Of these, even fewer (28.0%, or 7.6% of all caregivers) reported that this increase had been challenging for them (not shown).

Table 1 shows the distribution of caregivers and non-caregivers on independent variables. As shown, caregivers are generally slightly more likely than non-caregivers to be older, partnered, and non-employed, to have non-tertiary education, and to report poor health and poor social support. Patterns are quite similar for men and women and across the regions, except that the Oslo/Vestland sample is markedly younger and has more employed and higher education individuals. Of note, a minority of caregivers, and slightly more female (8−12%) than male (5−8%) caregivers, report being in poor health. A similar minority, but slightly more male (11−18%) than female (9−14%) caregivers, report poor social support. Furthermore, about one third of male caregivers and one fourth of female caregivers provide care to someone in their household, and between 23 and 30% of caregivers report that their caregiving responsibilities increased during the pandemic due to changes in the formal health and social services.

**Table 1:** Descriptive statistics (N (%)) of caregivers and non-caregivers

|  | Agder/Nordland (t1) |  |  |  | Oslo/Vestland (t3) |  |  |  |
| --- | --- | --- | --- | --- | --- | --- | --- | --- |
|  | Caregivers<br>(n=1,022) |  | Non-caregivers<br>(n=5,167) |  | Caregivers<br>(n=1,255) |  | Non-caregivers<br>(n=7,437) |  |
|  | Men<br>(n=493) | Women<br>(n=529) | Men<br>(n=2,452) | Women<br>(n=2,715) | Men<br>(n=548) | Women<br>(n=707) | Men<br>(n=3,291) | Women<br>(n=4,146) |
| Age 18–39 | 24 (4.9) | 32 (6.0) | 281 (11.5) | 544 (20.1) | 49 (9.0) | 87 (12.2) | 944 (28.7) | 1479 (35.6) |
| Age 40–59 | 243 (49.3) | 308 (58.1) | 1025 (41.8) | 1252 (46.1) | 306 (55.8) | 379 (53.7) | 1370 (41.6) | 1680 (40.5) |
| Age 60–92 | 226 (45.8) | 189 (35.7) | 1146 (46.8) | 919 (33.8) | 193 (35.3) | 241 (34.1) | 977 (29.7) | 987 (23.8) |
| Tertiary education | 232 (47.1) | 289 (54.6) | 1,294 (52.8) | 1,627 (59.9) | 343 (62.6) | 453 (64.1) | 2,217 (67.4) | 3,004 (72.5) |
| Partner | 437 (88.6) | 419 (79.2) | 2,081 (84.9) | 2,158 (79.5) | 462 (84.3) | 522 (73.8) | 2,633 (80.0) | 3,084 (73.5) |
| Employed | 314 (63.7) | 368 (69.6) | 1,576 (64.3) | 1,845 (68.0) | 405 (73.9) | 501 (70.9) | 2,546 (77.4) | 3,169 (76.4) |
| Poor health | 38 (7.7) | 63 (11.9) | 145 (5.9) | 202 (7.4) | 28 (5.1) | 57 (8.1) | 158 (4.8) | 256 (6.2) |
| Fair health | 103 (20.9) | 108 (20.4) | 370 (19.2) | 484 (17.8) | 100 (18.3) | 105 (14.9) | 488 (14.8) | 626 (15.1) |
| Good health | 352 (71.4) | 355 (67.1) | 1,834 (74.8) | 2,025 (74.6) | 419 (76.5) | 543 (76.8) | 2,633 (80.0) | 3,257 (78.6) |
| Poor social support | 55 (11.1) | 49 (9.3) | 218 (8.9) | 253 (9.3) | 101 (18.4) | 96 (13.6) | 547 (16.6) | 538 (13.0) |
| Moderate social support | 220 (44.6) | 226 (42.7) | 1,206 (49.2) | 1,154 (42.5) | 293 (53.5) | 327 (46.3) | 1,815 (55.2) | 2,095 (50.5) |
| Strong social support | 212 (43.0) | 251 (47.5) | 1,016 (41.4) | 1,292 (47.6) | 153 (27.9) | 277 (39.2) | 903 (27.4) | 1,497 (36.1) |
| Resident caregiving | 161 (32.7) | 132 (25.0) |  |  | 197 (36.0) | 179 (25.3) |  |  |
| Increased care load | 112 (22.7) | 136 (25.7) |  |  | 157 (28.7) | 214 (30.3) |  |  |

### 3.2 Change in psychosocial well-being

Figure 2 presents estimated levels of psychosocial well-being across the data points for caregivers and non-caregivers. First, it is evident that the overall trajectory of psychosocial well-being among all groups and across all outcomes is quite consistent: first characterized by stability, followed by a marked decline before plateauing or slightly improving in the later stage of the pandemic. As can also be seen, reported psychosocial well-being is generally lower in Oslo/Vestland than in Agder/Nordland.

Furthermore, psychological *well-being* tends to be slightly, but not significantly, higher among non-caregivers than caregivers both before and during the pandemic. This caregiver disadvantage is highest among men in Oslo/Vestland. Regarding *ill-being*, we see that caregivers reported higher levels than non-caregivers both before and during the pandemic. Women (irrespective of caregiver status) report higher levels of ill-being than men; in fact, men’s level during the peak of the pandemic is like that experienced by women before the pandemic. A similar pattern emerges also for *loneliness*, but only among men. Among women, change in loneliness is almost identical for caregivers and non-caregivers.

The most notable caregiver disadvantage is observed for ill-being, with caregivers reporting significantly higher levels of worry, anxiety, and sadness than their non-caregiving counterparts. The NCPHS includes also the 5-item Hopkins Symptom Checklist, which measures psychological distress (Strand et al., 2003). Analyzing change in this measure shows a similar pattern as that observed for ill-being (see online resource Figure S2), corroborating the plight of caregivers during COVID-19.

We observed similar results in sensitivity analyses including all available respondents (even if they only participated in one of the rounds). In these analyses caregivers were defined as giving care at least monthly at each given time point, meaning they could change from being caregivers and non-caregivers (see supplementary online Figure S3).

**Fig 2.**
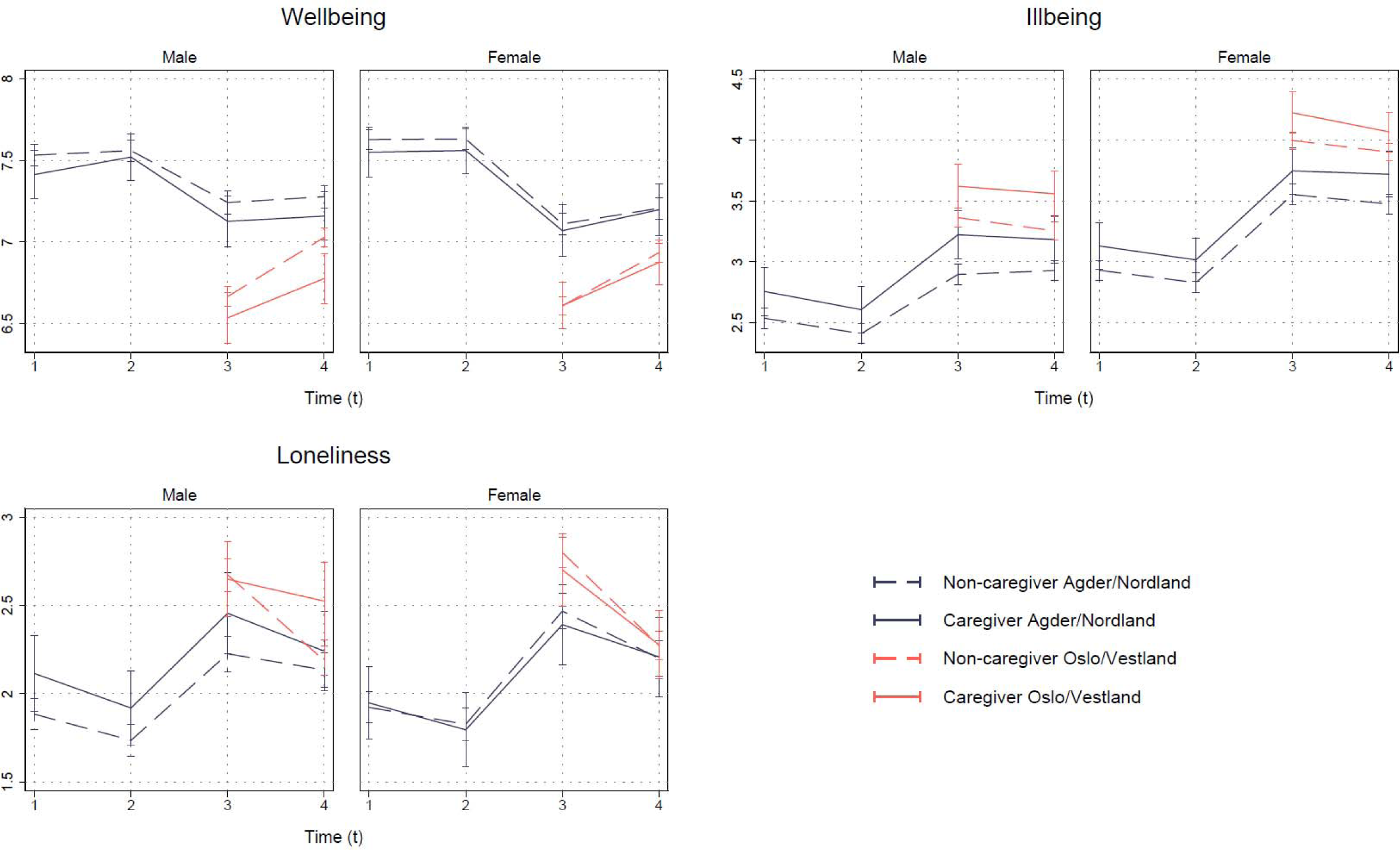
Trajectories of psychosocial indicators among caregivers and non-caregivers, adjusted for age, partner status, education, and employment status, and stratified by gender and county. Significant caregivers (vs. non-caregivers) (p< .05) associations are found only among males, for well-being/Oslo-Vestland/t4, ill-being/Agder-Nordland/t3, ill-being/Oslo-Vestland/both t3 and t4, and loneliness/Oslo-Vestland/t4 (see online supplementary material Table S1)

We have in auxiliary analyses (see supplementary Table S2) also analyzed the odds of reporting “low” well-being during the peak of the pandemic (controlling for sex, age, employment status, partner status, and education). We find that caregivers at t3 (Nov 2020) report significantly higher odds compared with non-caregivers of being worried (OR = 1.22, p< .01), anxious (OR = 1.23, p< .01), and depressed/sad (OR = 1.10, p< .05). Differences were not significant (p> .10) for happy (OR = 1.06), satisfied (OR = 1.13), or lonely (OR = 0.94). There were no significant sex differences in these results.

### 3.3 Predictors of caregiver psychosocial well-being during the pandemic

Table 2 shows the results of analyses of predictors of (change in) psychosocial well-being among caregivers. Patterns are generally quite similar for men and women. Older age, better health, and strong social support are associated with better psychosocial well-being across all indicators, although some of especially the longitudinal associations fail to reach statistical significance. Having a partner consistently relates to higher well-being and reduced loneliness but has no effect on ill-being. Educational level and employment status are mainly unrelated to all outcomes, except that being employed relates to slightly beneficial outcomes among women. These patterns of associations are quite similar among non-caregivers (see supplementary online resource Table S3). Turning to care-related factors, the analysis shows that residential caregiving (caring for someone within the household) is associated with slightly but not statistically significantly lower psychosocial well-being. Predictably, increased caregiving load during COVID-19 is associated with compromised psychosocial well-being along all indicators, also in longitudinal analyses.

We were interested in whether both trajectories and predictors of caregivers’ psychosocial well-being vary across age groups (supplementary online resources in Figure S4 and Table S4). We find no substantial age differences in the change in psychosocial well-being among caregivers. Similarly, age does not moderate associations between outcomes and caregiving factors (residential caregiving and increase caregiving load).

**Table 2:**
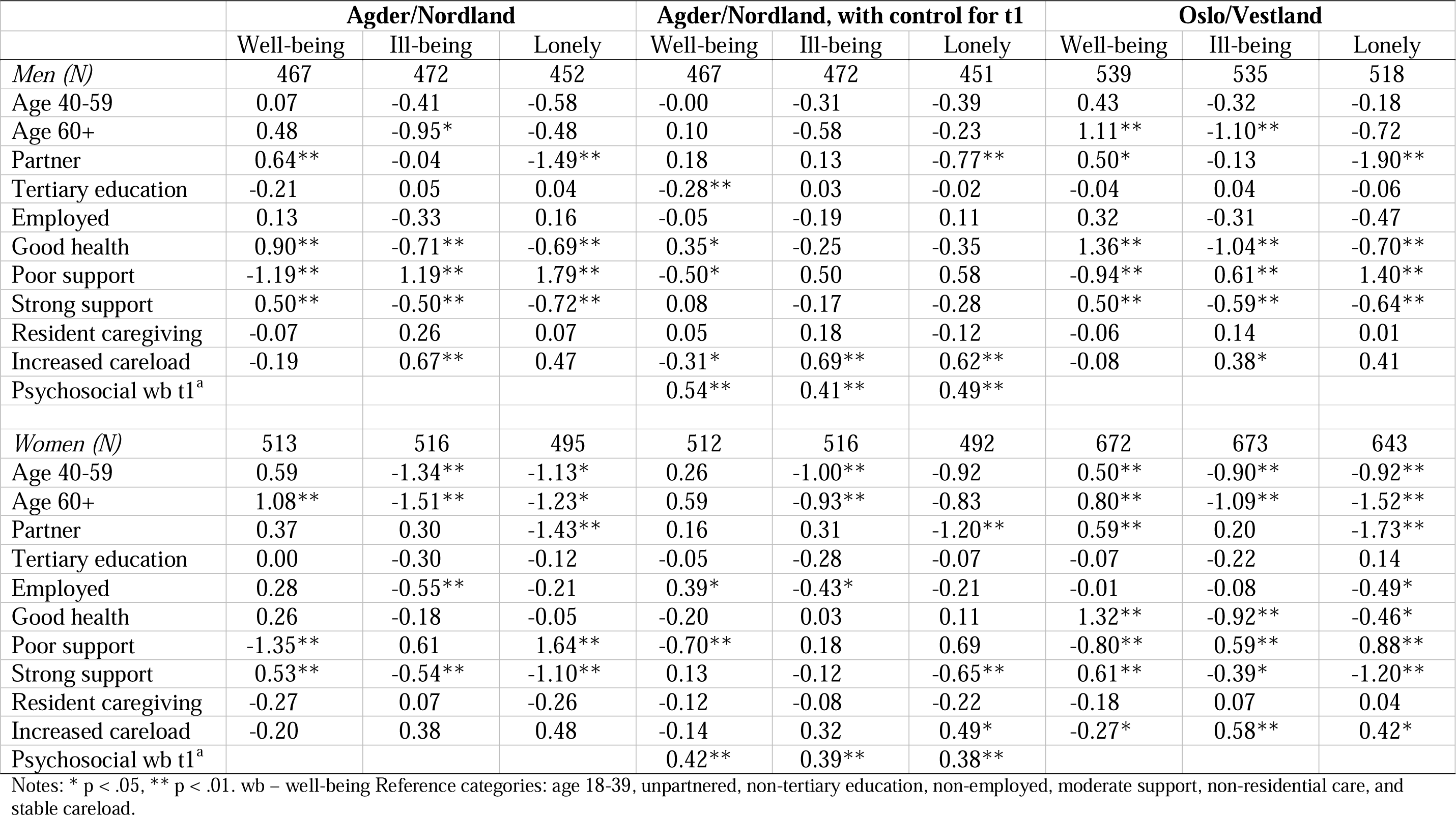
Regressing well-being indicators on background characteristics and care-related factors (data from t3/t4)

|  | **Agder/Nordland** |  |  | **Agder/Nordland, with control for t1** |  |  | **Oslo/Vestland** |  |  |
| --- | --- | --- | --- | --- | --- | --- | --- | --- | --- |
|  | Well-being | Ill-being | Lonely | Well-being | Ill-being | Lonely | Well-being | Ill-being | Lonely |
| *Men (N)* | 467 | 472 | 452 | 467 | 472 | 451 | 539 | 535 | 518 |
| Age 40-59 | 0.07 | -0.41 | -0.58 | -0.00 | -0.31 | -0.39 | 0.43 | -0.32 | -0.18 |
| Age 60+ | 0.48 | -0.95* | -0.48 | 0.10 | -0.58 | -0.23 | 1.11** | -1.10** | -0.72 |
| Partner | 0.64** | -0.04 | -1.49** | 0.18 | 0.13 | -0.77** | 0.50* | -0.13 | -1.90** |
| Tertiary education | -0.21 | 0.05 | 0.04 | -0.28** | 0.03 | -0.02 | -0.04 | 0.04 | -0.06 |
| Employed | 0.13 | -0.33 | 0.16 | -0.05 | -0.19 | 0.11 | 0.32 | -0.31 | -0.47 |
| Good health | 0.90** | -0.71** | -0.69** | 0.35* | -0.25 | -0.35 | 1.36** | -1.04** | -0.70** |
| Poor support | -1.19** | 1.19** | 1.79** | -0.50* | 0.50 | 0.58 | -0.94** | 0.61** | 1.40** |
| Strong support | 0.50** | -0.50** | -0.72** | 0.08 | -0.17 | -0.28 | 0.50** | -0.59** | -0.64** |
| Resident caregiving | -0.07 | 0.26 | 0.07 | 0.05 | 0.18 | -0.12 | -0.06 | 0.14 | 0.01 |
| Increased careload | -0.19 | 0.67** | 0.47 | -0.31* | 0.69** | 0.62** | -0.08 | 0.38* | 0.41 |
| Psychosocial wb t1<sup>a</sup> |  |  |  | 0.54** | 0.41** | 0.49** |  |  |  |
| *Women (N)* | 513 | 516 | 495 | 512 | 516 | 492 | 672 | 673 | 643 |
| Age 40-59 | 0.59 | -1.34** | -1.13* | 0.26 | -1.00** | -0.92 | 0.50** | -0.90** | -0.92** |
| Age 60+ | 1.08** | -1.51** | -1.23* | 0.59 | -0.93** | -0.83 | 0.80** | -1.09** | -1.52** |
| Partner | 0.37 | 0.30 | -1.43** | 0.16 | 0.31 | -1.20** | 0.59** | 0.20 | -1.73** |
| Tertiary education | 0.00 | -0.30 | -0.12 | -0.05 | -0.28 | -0.07 | -0.07 | -0.22 | 0.14 |
| Employed | 0.28 | -0.55** | -0.21 | 0.39* | -0.43* | -0.21 | -0.01 | -0.08 | -0.49* |
| Good health | 0.26 | -0.18 | -0.05 | -0.20 | 0.03 | 0.11 | 1.32** | -0.92** | -0.46* |
| Poor support | -1.35** | 0.61 | 1.64** | -0.70** | 0.18 | 0.69 | -0.80** | 0.59** | 0.88** |
| Strong support | 0.53** | -0.54** | -1.10** | 0.13 | -0.12 | -0.65** | 0.61** | -0.39* | -1.20** |
| Resident caregiving | -0.27 | 0.07 | -0.26 | -0.12 | -0.08 | -0.22 | -0.18 | 0.07 | 0.04 |
| Increased careload | -0.20 | 0.38 | 0.48 | -0.14 | 0.32 | 0.49* | -0.27* | 0.58** | 0.42* |
| Psychosocial wb t1<sup>a</sup> |  |  |  | 0.42** | 0.39** | 0.38** |  |  |  |

Notes: \* p < .05, \*\* p < .01. wb – well-being Reference categories: age 18-39, unpartnered, non-tertiary education, non-employed, moderate support, non-residential care, and stable careload.

## 4. Discussion

To our knowledge, this is the first longitudinal study of trajectories in caregivers’ psychosocial well-being from before and during different stages of the COVID-19 pandemic. Our study demonstrates that levels of psychosocial well-being first remained stable but later dropped markedly during the peak stages of the pandemic. With some variations, and in line with previous research, caregivers report lower levels compared with non-caregivers both before and during the pandemic. While the magnitude of the declines in psychosocial well-being is similar across the two groups, the declines affect caregivers more: “a falling tide sinks all boats”, yet the implications are graver for those lower on the well-being ladder.

Regarding psychological ill-being (i.e., negative emotionality), caregivers fare worse than non-caregivers both before and during the pandemic. However, caregivers are at higher risk of reporting severe emotional reactions as indicated by high levels of worry, anxiety, or sadness during the pandemic. These findings echo those of previous cross-sectional studies from the beginning of the pandemic, documenting increased caregiver burden and distress (Altieri and Santangelo 2021; Canevelli et al. 2020; Cohen et al. 2021; Truskinovsky et al. 2022). In contrast to this literature, which finds more serious impacts among women, we find similar impacts for men and women. This difference likely reflects the relatively high access to formal care and gender equality in caregiving roles in Norway, mitigating risks for female caregivers in particular.

Against a background literature which portrays caregiving as detrimental for loneliness and well-being (e.g., Pinquart and Sörensen 2006; Vasileiou et al. 2017), it is noteworthy that this and prior Norwegian studies (Hansen and Slagsvold 2015; Hansen et al. 2013) find no such overall impacts. Across all time points, caregivers report similar levels of evaluative (life satisfaction) and experienced (joy) well-being compared with non-caregivers. Our findings are consistent with notions that caregiving, including in the COVID-era, may confer gains as well as strains (Hansen and Slagsvold 2012). As noted, patterns likely also mirror that caregiving is relatively less demanding in Norway and other countries with comprehensive formal care services and generous social welfare protections (Colombo 2011; Hansen et al. 2013). Similarly, Norway had relatively low pandemic-related infection and mortality rates, potentially explaining fewer COVID-era psychological impacts than in other countries. These notions are supported by the fact that few (28%) caregivers in our sample report that their caregiving load increased due to changes in the health- and social services during the peak of the pandemic, of which less than one third reported the changes as “challenging”.

The somewhat surprising finding that caregiving seems inconsequential for men and women’s cognitive well-being, even during lockdown when caregiving can be extra challenging, may attest to the highly cognitive nature of satisfaction judgments. These appraisals may be detached from, or even enhanced by, emotionally taxing and burdensome experiences (Hansen 2010). The near-zero effects also suggest that, although aspects of caregiving may reduce satisfaction and joy, other aspects (e.g., helping others, feeling useful and needed, receiving appraisal) may promote positive self-evaluations. Similar predictions can be made also regarding loneliness. While caregiving can limit engagement in social activities, it can also, especially during a time of crisis and isolation, improve a sense of continuity, social connection, and purpose, potentially helping to stave off loneliness. Finally, the near-zero findings likely mirror that reactions are highly heterogenous: some caregivers may be frustrated by limitations on daily routines and access to formal supports, others experienced few major changes to their already solitary and home-based lifestyle (Savla et al. 2021).

Which subgroups of caregivers were the most vulnerable? We assessed a handful of potential risk and protective factors for negative (change in) psychosocial well-being during the peak of the pandemic. Key risk factors are younger age, poorer health, being unpartnered, lacking social support, resident (in-household) caregiving, and experiencing more care responsibility during the pandemic, although some of the longitudinal associations fail to reach statistical significance. Of note, the analysis highlights the importance of social support. A sizeable percentage of caregivers (10–15%), and more among those with low psychosocial well-being, report having poor access to social support during the pandemic. A similar percentage of caregivers report health problems themselves, and may find caregiving particularly challenging due to their own COVID-19 exposure risks (MacLeod et al. 2021). Another noteworthy finding is the lack of a social gradient in caregivers’ psychosocial well-being, again potentially reflecting the idea of equal access to health services under the Norwegian welfare system. As expected, caregiver distress is higher among those providing the most intensive care—that is, to someone in the household, and among those experiencing increasing care responsibility during the pandemic.

This study has several strengths, most notably within-person data from different stages of the pandemic, the scope of variables, and the large sample size, providing rich possibilities for capturing the complexities of caregiving, over time, and for different subgroups. There are some weaknesses to note. For example, we lack details about type of care (e.g., personal care vs. instrumental care), relation to the care recipient, and the care recipient’s health. We are thus unable to do more subgroup analyses of caregivers, or to assess whether it is caregiving in itself or deteriorating health of a close relative that is most impactful. Over the study period of up to 27 months, many care recipients are likely to experience increased health problems with negative psychological implications, irrespective of the pandemic, also for their caregiver. Furthermore, there are potential weaknesses related to the use of single-item measures and unvalidated scales. Although our individual single-item measures are commonly used and recommended measurements in the field, the composite indexes should be validated in future research. First, findings should be interpreted in light of Norway’s relatively low COVID-19 morbidity and mortality rates and relaxed infection control measures. Of note, our two countries with pre-pandemic data (Agder and Nordland) were among the counties least impacted by COVID-19 in Norway. Coupled with the comparatively extensive welfare supports in Norway, pandemic-related caregiver distress is likely greater in other countries.

To conclude, while psychosocial well-being declined overall during the pandemic, the impacts seem somewhat more serious for caregivers who reported lower psychosocial well-being already before the pandemic. The psychosocial costs of COVID-19 for caregivers were especially notable in terms of elevated negative emotions such as worries and distress. These costs were also more pronounced in urban regions with more pandemic-related infections and restrictions, and among caregivers who provide more intensive care, have their own health problems, or lack social support. That said, the impacts seem milder than suggested by prior research (e.g., Eurocarers 2021). The contrast could reflect that our within-person design avoids some of the bias in earlier cross-sectional and retrospective subjective data (e.g., recall bias). It could also stem from country differences, thus highlighting the need to replicate our analysis in countries with fewer social protections, a greater age-specific disease burden, higher mortality rates, and more stringent social restrictions during the pandemic.

Understanding how caregivers reacted to the difficulties imposed by the pandemic is essential to support at-risk caregivers, and, by extension, their care recipients, during future pandemics or times of crisis. It is also key to future balancing of health protective measures against their unintended consequences for the well-being and health of vulnerable groups. The importance is highlighted also by the well-established consequences of compromised psychosocial well-being on physical and mental health (e.g., Steptoe et al. 2013). These effects in turn impact on their ability to provide care and increase the risk of institutionalization and additional health and social costs (Gallagher and Wetherell 2020). Attempts to reduce caregiver burden, especially during times of crisis, thus has clear implications for the health and functioning of people in and around the care relationships, as well as for wider society.

## Data Availability

Data are available upon application to www.helsedata.no

## Footnotes

1 Respondents in Oslo and Vestland were asked retrospectively only about Nov−Dec 2020.

## References

Abbas, J. (2021). Crisis management, transnational healthcare challenges and opportunities: The intersection of COVID-19 pandemic and global mental health. Research in Globalization, 100037. https://doi.org/10.1016/j.resglo.2021.100037

Adelman, R. D., Tmanova, L. L., Delgado, D., Dion, S., and Lachs, M. S. (2014). Caregiver burden: a clinical review. JAMA, 311(10), 1052–1060.

Altieri, M., and Santangelo, G. (2021). The psychological impact of COVID-19 pandemic and lockdown on caregivers of people with dementia. The American Journal of Geriatric Psychiatry, 29(1), 27–34.

Blake-Holmes, K., and McGowan, A. (2022). ‘It’s making his bad days into my bad days’: The impact of coronavirus social distancing measures on young carers and young adult carers in the United Kingdom. Child and Family Social Work, 27(1), 22–29.

Bøen, H., Dalgard, O. S., and Bjertness, E. (2012). The importance of social support in the associations between psychological distress and somatic health problems and socio-economic factors among older adults living at home: a cross sectional study. BMC geriatrics, 12(1), 1–12.

Canevelli, M., Valletta, M., Blasi, M. T., Remoli, G., Sarti, G., Nuti, F., . . . Bruno, G. (2020). Facing dementia during the COVID[19 outbreak. Journal of the American Geriatrics Society, 68(8), 1673–1676.

Cohen, S. A., Kunicki, Z. J., Drohan, M. M., and Greaney, M. L. (2021). Exploring changes in caregiver burden and caregiving intensity due to COVID-19. Gerontology and Geriatric Medicine, 7, 2333721421999279.

Colombo, F. (2011). Help Wanted? Providing and Paying for Long-Term Care, OECD Health Policy Studies, OECD Publishing.

Diener, E. (2012). New findings and future directions for subjective well-being research. American Psychologist, 67(8), 590.

Daatland, S. O., Herlofson, K., and Lima, I. A. (2011). Balancing generations: on the strength and character of family norms in the West and East of Europe. Ageing and Society, 31(7), 1159–1179.

Eurocarers. (2021). Impact of the COVID-19 outbreak on informal carers across Europe - Final report. Retrieved from Brussels/Ancona:

Eurofound. (2022). COVID-19 and older people: impact on their lives, support and care (9289722444). Publications Office of the European Union, Luxembourg.

Gallagher, S., and Wetherell, M. A. (2020). Risk of depression in family caregivers: unintended consequence of COVID-19. BJPsych open, 6(6).

Gaugler, J. E., Kane, R. L., and Newcomer, R. (2007). Resilience and transitions from dementia caregiving. The Journals of Gerontology Series B: Psychological Sciences and Social Sciences, 62(1), P38–P44.

Hansen, T. (2010). Subjective well-being in the second half of life: The influence of family and household resources. PhD thesis. University of Oslo.

Hansen, T., Nilsen, T. S., Yu, B., Knapstad, M., Skogen, J. C., Vedaa, Ø., and Nes, R. B. (2021). Locked and lonely? A longitudinal assessment of loneliness before and during the COVID-19 pandemic in Norway. Scandinavian Journal of Public Health, 1403494821993711.

Hansen, T., and Slagsvold, B. (2012). The Strains and Gains of Caregiving: An Examination of the Effects of Providing Personal Care to a Partner on a Range of Psychological Outcomes. Gerontologist, 52, 515–516.

Hansen, T., and Slagsvold, B. (2015). Feeling the squeeze? The effects of combining work and informal caregiving on psychological well-being. European Journal of Ageing, 12(1), 51–60. doi:10.1007/s10433-014-0315-y

Hansen, T., Slagsvold, B., and Ingebretsen, R. (2013). The Strains and Gains of Caregiving: An Examination of the Effects of Providing Personal Care to a Parent on a Range of Indicators of Psychological Well-Being. Social Indicators Research, 114(2), 323–343. doi:10.1007/s11205-012-0148-z

Kent, E. E., Ornstein, K. A., and Dionne-Odom, J. N. (2020). The family caregiving crisis meets an actual pandemic. Journal of pain and symptom management, 60(1), e66–e69.

Leggett, A., Koo, H. J., Park, B., and Choi, H. J. (2022). The Changing Tides of Caregiving During the COVID-19 Pandemic: How Decreasing and Increasing Care Provision Relates to Caregiver Well-Being. The Journals of Gerontology: Series B.

Lightfoot, E., Yun, H., Moone, R., Otis, J., Suleiman, K., Turck, K., and Kutzler, C. (2021). Changes to family caregiving of older adults and adults with disabilities during COVID-19. Gerontology and Geriatric Medicine, 7, 23337214211002404.

MacLeod, S., Tkatch, R., Kraemer, S., Fellows, A., McGinn, M., Schaeffer, J., and Yeh, C. S. (2021). The impact of COVID-19 on informal caregivers in the US. International Journal of Aging Research, 4(3), 87–87.

Meltzer, H. (2003). Development of a common instrument for mental Health In: Nosikov A, Gudex C (eds) EUROHIS: developing common istruments for health surveys. Amsterdam: IOS Press.

Merlani, P., Verdon, M., Businger, A., Domenighetti, G., Pargger, H., and Ricou, B. (2011). Burnout in ICU caregivers: a multicenter study of factors associated to centers. American Journal of Respiratory and Critical Care Medicine, 184(10), 1140-1146.

Nes, R. B., Hansen, T., and Barstad, A. (2018). Livskvalitet: Anbefalinger for et bedre målesystem [Quality of life: Measurement guidelines]. Oslo: The National Health Directorate.

Nørgaard, S. K., Vestergaard, L. S., Nielsen, J., Richter, L., Schmid, D., Bustos, N., . . . Denissov, G. (2021). Real-time monitoring shows substantial excess all-cause mortality during second wave of COVID-19 in Europe, October to December 2020. Eurosurveillance, 26(2), 2002023.

OECD. (2013). OECD guidelines on measuring subjective well-being. OECD Publishing.

Onwumere, J., Creswell, C., Livingston, G., Shiers, D., Tchanturia, K., Charman, T., . . . Wildman, E. (2021). COVID-19 and UK family carers: policy implications. The Lancet Psychiatry, 8(10), 929-936.

Opinion. (2021). Nasjonal pårørendeundersøkelse [National carer survey]. Oslo: Opinion.

Park, S. S. (2021). Caregivers’ mental health and somatic symptoms during COVID-19. The Journals of Gerontology: Series B, 76(4), e235-e240.

Pinquart, M., and Sörensen, S. (2006). Gender differences in caregiver stressors, social resources, and health: An updated meta-analysis. The Journals of Gerontology Series B: Psychological Sciences and Social Sciences, 61(1), P33–P45.

Savla, J., Roberto, K. A., Blieszner, R., McCann, B. R., Hoyt, E., and Knight, A. L. (2021). Dementia caregiving during the “stay-at-home” phase of COVID-19 pandemic. The Journals of Gerontology: Series B, 76(4), e241–e245.

Skirbekk, V., Dieleman, J. L., Stonawski, M., Fejkiel, K., Tyrovolas, S., and Chang, A. Y. (2022). The health-adjusted dependency ratio as a new global measure of the burden of ageing: a population-based study. The Lancet Healthy Longevity, 3(5), e332–e338.

Steptoe, A., Shankar, A., Demakakos, P., and Wardle, J. (2013). Social isolation, loneliness, and all-cause mortality in older men and women. Proceedings of the National Academy of Sciences, 110(15), 5797–5801.

Strand, B. H., Dalgard, O. S., Tambs, K., and Rognerud, M. (2003). Measuring the mental health status of the Norwegian population: A comparison of the instruments SCL-25, SCL-10, SCL-5 and MHI-5 (SF-36). Nordic Journal of Psychiatry, 57(2), 113-118. doi:10.1080/08039480310000932

The Norwegian Government. (2022). Timeline: News from Norwegian Ministries about the Coronavirus disease Covid-19. Retrieved June 10 2022 from https://www.regjeringen.no/en/topics/koronavirus-covid-19/timeline-for-news-from-norwegian-ministries-about-the-coronavirus-disease-covid-19/id2692402/

Truskinovsky, Y., Finlay, J. M., and Kobayashi, L. C. (2022). Caregiving in a Pandemic: COVID-19 and the Well-Being of Family Caregivers 55+ in the United States. Medical Care Research and Review, 10775587211062405.

Vasileiou, K., Barnett, J., Barreto, M., Vines, J., Atkinson, M., Lawson, S., and Wilson, M. (2017). Experiences of loneliness associated with being an informal caregiver: a qualitative investigation. Frontiers in psychology, 8, 585.

WHO. (2022). WHO coronavirus disease (COVID-19) - Norway situation. Retrieved June 1 2022 from https://covid19.who.int/region/euro/country/no

Zwar, L., König, H.-H., and Hajek, A. (2021). Informal caregiving during the COVID-19 pandemic: findings from a representative, population-based study during the second wave of the pandemic in Germany. Aging and Mental Health, 1-9.

Aartsen, M., and Hansen, T. (2020). Social Participation in the Second Half of Life. In S. Rattan (Ed.), Encyclopedia of Biomedical Gerontology (Vol. 247-255). London: Academic Press. https://doi.org/10.1016/b978-0-12-801238-3.11351-0

