## Supplementary files for "Emerging costs in a “hidden” workforce: The longitudinal psychosocial effects of caregiving during the COVID-19 pandemic among Norwegian adults"

By Hansen et al, 2023.

(Short titles)


| | |
|---|---|
| **Supplementary Fig 1:** Transition plot. | p. 2 |
| **Supplementary Fig 2:** Hopkins Symptom Checklist as outcome. | p. 3 |
| **Supplementary Fig 3:** Trajectories including full sample. | p. 4 |
| **Supplementary Table 1:** Significance tests for figure 2. | p. 4 |
| **Supplementary Table 2:** Low values of wellbeing. | p. 5 |
| **Supplementary Table 3:** Predictors of well-being among non-caregivers. | p. 5 |
| **Supplementary Fig 4:** Age and wellbeing. | p. 6 |
| **Supplementary Table 4:** Wellbeing predictors for various age groups. | p. 7 |

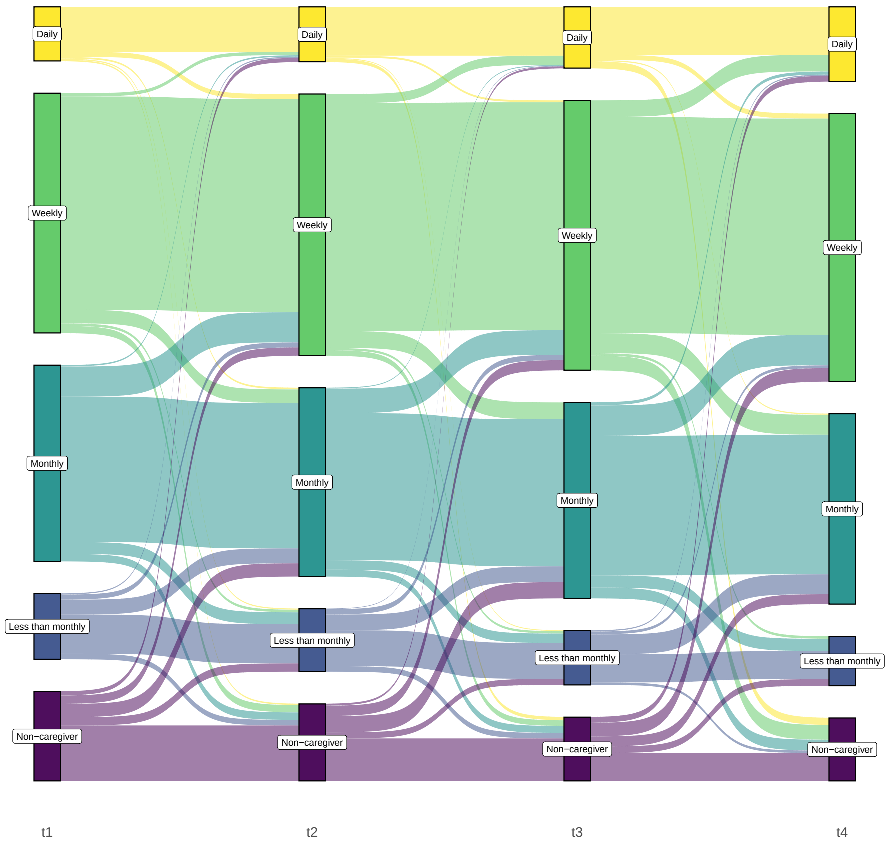


Note: Figure shows the share of caregivers providing care at least daily (yellow), weekly (green), and monthly (turquoise), less than monthly (blue) and non-caregivers (purple) at the four time points. Only those providing care at at least one time point were included in the figure. The figure also shows the flow – how caregivers move from providing care e.g. daily to e.g. weekly between time points. Only persons from Nordland and Agder were included.

**Supplementary Fig 1:** Transition plot: How persons move through caregiving roles.

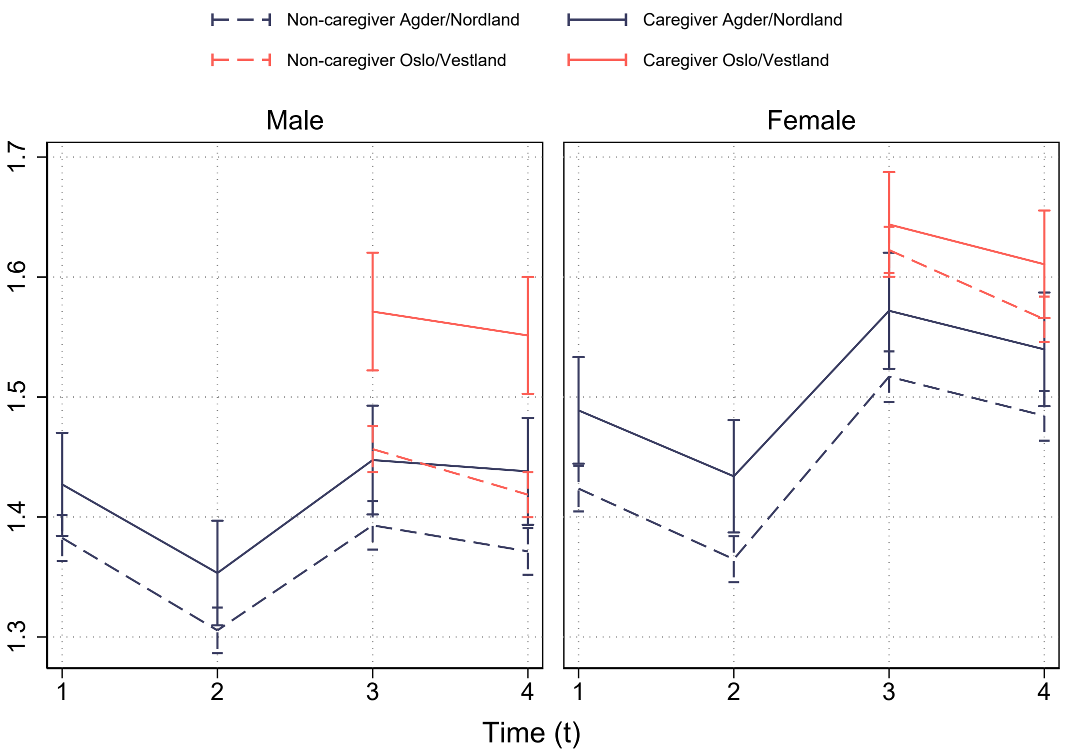


**Supplementary Fig 2:** Hopkins Symptom Checklist as outcome.

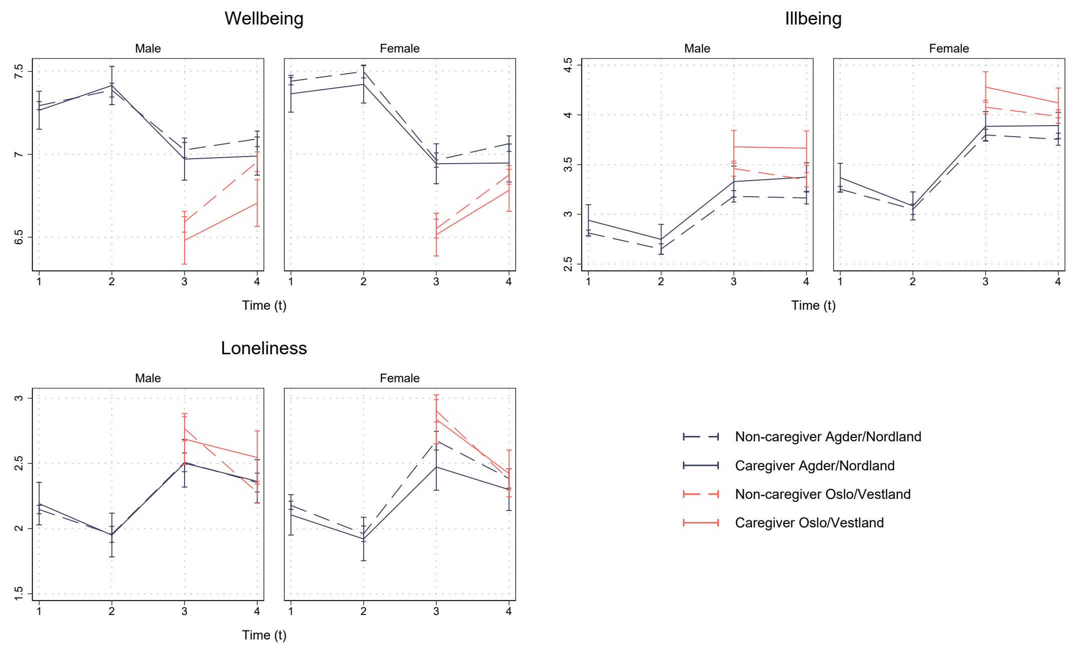


Note: Sensitivity analysis including all 60.000 who participated in at least one of the surveys. In this analysis persons are also allowed to move between being caregivers and non-caregivers.

**Supplementary Fig 3:** Trajectories including the full sample (n = 60 000).

**Supplementary Table 1:** Significance tests for figure 2.

Significance between caregivers and non-caregivers from figure 2.

| | | Wellbeing | | Illbeing | | Loneliness | |
|---|---|---|---|---|---|---|---|
| | | Male | Female | Male | Female | Male | Female |
| AGDER/NORDLAND | | | | | | | |
| t1 | Caregiver | 7.41 (7.27-7.56) | 7.55 (7.40-7.70) | 2.76 (2.56-2.96) | 3.13 (2.94-3.32) | 2.11 (1.90-2.33) | 1.95 (1.74-2.15) |
| | Non-caregiver | 7.53 (7.47-7.60) | 7.63 (7.57-7.69) | 2.54 (2.45-2.62) | 2.93 (2.85-3.01) | 1.88 (1.80-1.97) | 1.92 (1.83-2.01) |
| t2 | Caregiver | 7.52 (7.38-7.67) | 7.56 (7.42-7.71) | 2.61 (2.42-2.80) | 3.02 (2.84-3.19) | 1.92 (1.71-2.13) | 1.80 (1.59-2.00) |
| | Non-caregiver | 7.56 (7.50-7.63) | 7.63 (7.57-7.69) | 2.41 (2.33-2.49) | 2.83 (2.75-2.91) | 1.74 (1.64-1.83) | 1.83 (1.73-1.92) |
| t3 | Caregiver | 7.13 (6.97-7.28) | 7.07 (6.91-7.23) | 3.22 (3.02-3.42)* | 3.75 (3.56-3.94) | 2.46 (2.22-2.69) | 2.39 (2.16-2.62) |
| | Non-caregiver | 7.24 (7.17-7.31) | 7.11 (7.04-7.18) | 2.90 (2.81-2.98) | 3.55 (3.47-3.64) | 2.23 (2.13-2.33) | 2.47 (2.37-2.57) |
| t4 | Caregiver | 7.16 (7.01-7.31) | 7.20 (7.04-7.36) | 3.18 (2.99-3.37) | 3.72 (3.53-3.90) | 2.24 (2.01-2.47) | 2.21 (1.98-2.43) |
| | Non-caregiver | 7.28 (7.21-7.35) | 7.21 (7.14-7.27) | 2.93 (2.85-3.01) | 3.47 (3.39-3.56) | 2.13 (2.04-2.23) | 2.20 (2.10-2.30) |
| OSLO/VESTLAND | | | | | | | |
| t3 | Caregiver | 6.53 (6.38-6.69) | 6.61 (6.47-6.75) | 3.62 (3.44-3.80)* | 4.22 (4.05-4.39) | 2.65 (2.44-2.86) | 2.69 (2.49-2.90) |
| | Non-caregiver | 6.67 (6.60-6.73) | 6.61 (6.55-6.67) | 3.36 (3.29-3.44) | 4.00 (3.93-4.07) | 2.67 (2.58-2.77) | 2.80 (2.71-2.89) |
| t4 | Caregiver | 6.78 (6.62-6.93)* | 6.88 (6.74-7.01) | 3.56 (3.37-3.74)* | 4.07 (3.90-4.23) | 2.52 (2.30-2.74)* | 2.28 (2.08-2.47) |
| | Non-caregiver | 7.03 (6.97-7.09) | 6.93 (6.88-6.99) | 3.26 (3.18-3.33) | 3.90 (3.83-3.97) | 2.19 (2.10-2.27) | 2.27 (2.19-2.35) |

* $p < 0.05$

**Supplementary Table 2:** Low values of wellbeing.

Table shows the odds ratio for caregivers vs. non-caregivers for low values on wellbeing items where low values are characterised as lower than 4, 3 or 2. Here we have changed the scale for negative worded items (e.g. for lonely$\leq$ 4 would be the same as $\geq$ 6).

| | $\leq 4$ | $\leq 3$ | $\leq 2$ |
|---|---|---|---|
| Happy | 1.06 | 1.19** | 1.23 |
| Satisfied | 1.13 | 1.18* | 1.64*** |
| Worried | 1.22*** | 1.23*** | 1.27*** |
| Down and sad | 1.10* | 1.17** | 1.25** |
| Anxious | 1.23*** | 1.22*** | 1.20** |
| Lonely | 0.94 | 0.86** | 0.91 |

* $p < 0.1$, ** $p < 0.05$, *** $p < 0.01$

**Supplementary Table 3:** Predictors of well-being among non-caregivers.

| | Agder/Nordland | | | Agder/Nordland with lag | | | Oslo/Vestland | | |
|---|---|---|---|---|---|---|---|---|---|
| | Wellbeing | Illbeing | Loneliness | Wellbeing | Illbeing | Loneliness | Wellbeing | Illbeing | Loneliness |
| **Men (N)** | 2.363 | 2.361 | 2.282 | 2.360 | 2.359 | 2.281 | 3.176 | 3.195 | 3.107 |
| Age 40-59 | 0.26** | −0.47*** | −0.42*** | 0.08 | −0.39*** | −0.25* | 0.24*** | −0.53*** | −0.52*** |
| Age 60+ | 0.60*** | −0.88*** | −0.53*** | 0.16* | −0.57*** | −0.23 | 0.70*** | −1.14*** | −0.75*** |
| Partner | 0.21** | 0.00 | −1.84*** | −0.04 | 0.09 | −1.04*** | 0.55*** | −0.15* | −2.28*** |
| Tertiary education | −0.11* | −0.01 | 0.00 | 0.01 | 0.00 | 0.03 | −0.07 | 0.11 | 0.12 |
| Employed | 0.08 | −0.15 | −0.10 | 0.01 | −0.09 | −0.10 | 0.02 | −0.17* | −0.15 |
| Good health | 0.93*** | −1.00*** | −0.63*** | 0.32*** | −0.57*** | −0.32*** | 1.22*** | −1.32*** | −0.90*** |
| Poor support | −0.85*** | 0.69*** | 1.04*** | −0.12 | 0.17 | 0.32* | −0.67*** | 0.38*** | 0.97*** |
| Strong support | 0.57*** | −0.46*** | −0.70*** | 0.17*** | −0.15** | −0.26*** | 0.55*** | −0.45*** | −0.74*** |
| Psychosocial wb t1[a] | | | | 0.55*** | 0.41*** | 0.46*** | | | |
| **Women (N)** | 2.593 | 2.613 | 2.531 | 2.582 | 2.609 | 2.526 | 4.019 | 4.047 | 3.921 |
| Age 40-59 | 0.20** | −0.65*** | −0.58*** | 0.06 | −0.43*** | −0.39*** | 0.12** | −0.77*** | −0.54*** |
| Age 60+ | 0.43*** | −0.72*** | −0.52*** | 0.02 | −0.30*** | −0.20 | 0.39*** | −1.05*** | −0.83*** |
| Partner | 0.51*** | −0.17* | −1.54*** | 0.21*** | −0.04 | −0.99*** | 0.66*** | −0.21*** | −2.05*** |
| Tertiary education | −0.09 | −0.09 | −0.02 | 0.03 | −0.04 | −0.08 | −0.23*** | 0.07 | 0.20** |
| Employed | 0.10 | −0.06 | −0.12 | 0.04 | −0.02 | −0.15 | 0.01 | −0.04 | −0.24** |
| Good health | 0.87*** | −0.69*** | −0.70*** | 0.26*** | −0.29*** | −0.32*** | 1.23*** | −1.14*** | −0.94*** |
| Poor support | −0.85*** | 0.63*** | 1.24*** | −0.14 | 0.09 | 0.24 | −0.93*** | 0.62*** | 1.35*** |
| Strong support | 0.49*** | −0.41*** | −0.97*** | 0.13** | −0.09 | −0.54*** | 0.56*** | −0.42*** | −0.89*** |
| Psychosocial wb t1[a] | | | | 0.53*** | 0.44*** | 0.46*** | | | |

* $p < 0.1$, ** $p < 0.05$, *** $p < 0.01$

[a] respective indicator (e.g. well-being at t1 for the first column, ill-being at t1 for the second, etc.).

### **Supplementary Fig 4:** Age and wellbeing.

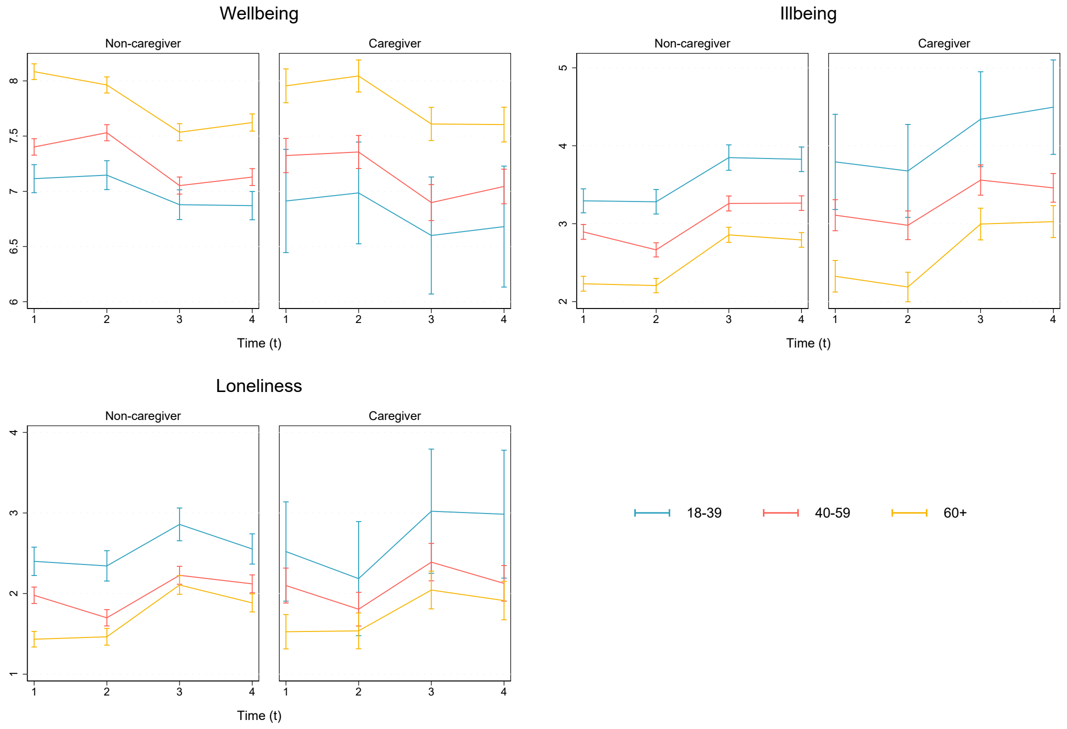


Note: Figure shows well-being for the various age groups, stratified by caregiver status. Estimates are adjusted for partner status, education, and employment status. To make graphs less noisy only Nordland and Agder were included.

**Supplementary Table 4:** Wellbeing predictors for caregivers divided by age groups.

A

| | Wellbeing | | | | | |
|---|---|---|---|---|---|---|
| | *18-29* | *30-39* | *40-49* | *50-59* | *60-69* | *70+* |
| Female | 0.08 | -0.24 | 0.10 | -0.07 | -0.17 | -0.23 |
| Partner | -0.15 | 0.32 | 0.85*** | 0.71*** | 0.40** | -0.15 |
| Higher ed | -0.08 | 0.10 | -0.15 | -0.04 | -0.21* | -0.48** |
| Employed | 0.94 | -0.36 | 0.38 | 0.15 | 0.06 | 0.33 |
| Good health | 0.31 | 1.59*** | 0.66*** | 0.76*** | 1.00*** | 1.21*** |
| Poor support | -1.65** | -0.92** | -1.57*** | -1.04*** | -0.67** | -0.28 |
| Strong support | 0.54 | 0.31 | 0.36** | 0.82*** | 0.49*** | 0.51** |
| **Care related** | | | | | | |
| Residential | -0.78* | 0.37 | -0.47** | 0.13 | -0.13 | -0.45** |
| Increased | -0.29 | -0.64* | -0.03 | -0.27** | -0.17 | -0.05 |
| Constant | 5.41 | 5.86 | 5.23 | 5.69 | 6.54 | 7.10 |
| Observations | 61 | 125 | 406 | 808 | 605 | 188 |

B

| | Illbeing | | | | | |
|---|---|---|---|---|---|---|
| | *18-29* | *30-39* | *40-49* | *50-59* | *60-69* | *70+* |
| Female | 1.01* | 0.99** | 0.31 | 0.51*** | 1.05*** | 0.83*** |
| Partner | 1.53*** | -0.63 | -0.23 | 0.10 | 0.35* | 0.37 |
| Higher ed | -0.39 | -0.63 | -0.01 | 0.11 | -0.21 | 0.03 |
| Employed | -1.41** | 0.32 | -0.35 | -0.36 | -0.20 | -0.16 |
| Good health | -0.14 | -0.70* | -0.50** | -0.63*** | -0.82*** | -0.66* |
| Poor support | 1.12 | 0.97** | 1.21*** | 0.55** | 0.80*** | -0.23 |
| Strong support | -0.02 | -0.49 | -0.38* | -0.85*** | -0.27* | -0.70** |
| **Care related** | | | | | | |
| Residential | 0.21 | -0.34 | 0.16 | -0.02 | 0.31* | 0.21 |
| Increased | 0.83 | 1.01** | 0.21 | 0.49*** | 0.61*** | 0.39 |
| Constant | 3.09 | 3.76 | 4.21 | 3.51 | 2.02 | 2.50 |
| Observations | 61 | 126 | 409 | 813 | 604 | 185 |

C

| | Loneliness | | | | | |
|---|---|---|---|---|---|---|
| | *18-29* | *30-39* | *40-49* | *50-59* | *60-69* | *70+* |
| Female | 0.51 | 0.86** | 0.42* | -0.11 | 0.09 | 0.17 |
| Partner | -1.41* | -1.54*** | -2.19*** | -1.58*** | -1.56*** | -0.48 |
| Higher ed | 0.14 | -0.11 | -0.07 | 0.01 | 0.04 | 0.23 |
| Employed | -1.49* | 0.78 | -0.68* | -0.42 | -0.03 | -0.39 |
| Good health | 0.73 | -1.27** | -0.53* | 0.00 | -0.61*** | -1.24** |
| Poor support | 0.56 | 2.52*** | 1.66*** | 1.53*** | 0.71** | 0.34 |
| Strong support | -1.57** | -1.03** | -0.92*** | -1.11*** | -0.74*** | -0.79** |
| **Care related** | | | | | | |
| Residential | 1.48** | -0.42 | -0.07 | -0.31* | 0.10 | 0.20 |
| Increased | 1.53** | -0.02 | -0.04 | 0.60*** | 0.72*** | -0.50 |
| Constant | 4.14 | 3.11 | 4.74 | 4.28 | 3.67 | 3.36 |
| Observations | 61 | 124 | 394 | 785 | 581 | 165 |
